# From “don, doff, and discard” to “don, doff, and decontaminate” – determination of filtering facepiece respirator and surgical mask integrity and inactivation of a SARS-CoV-2 surrogate and a small non-enveloped virus following multiple-cycles of vaporised hydrogen peroxide, ultraviolet germicidal irradiation, and dry heat decontamination

**DOI:** 10.1101/2021.01.15.21249866

**Authors:** Louisa F. Ludwig-Begall, Constance Wielick, Olivier Jolois, Lorène Dams, Ravo M. Razafimahefa, Hans Nauwynck, Pierre-Francois Demeuldre, Aurore Napp, Jan Laperre, Frédéric Farnir, Etienne Thiry, Eric Haubruge

**Affiliations:** Veterinary Virology and Animal Viral Diseases, Department of Infectious and Parasitic Diseases, FARAH Research Centre, Faculty of Veterinary Medicine, Liège University, Liège, Belgium; Centexbel Textile Research Centre, Grace-Hollogne, Belgium; Laboratory of Virology, Faculty of Veterinary Medicine, Ghent University, Merelbeke, Belgium; Department of Hospital Pharmacy, The University Hospital Center, Liège University, Liège, Belgium; Biostatistics and Bioinformatics Applied to Veterinary Science, FARAH Research Centre, Faculty of Veterinary Medicine, University of Liège, Liège, Belgium; TERRA Research Centre, Gembloux Agro-Bio Tech, Liège University, Gembloux, Belgium

## Abstract

**Background:** As the SARS-CoV-2 pandemic accelerates, the supply of personal protective equipment remains under strain. To combat shortages, re-use of surgical masks and filtering facepiece respirators has been recommended. Prior decontamination is paramount to the re-use of these typically single-use only items and, without compromising their integrity, must guarantee inactivation of SARS-CoV-2 and other contaminating pathogens.

**Aim:** We provide information on the effect of time-dependent passive decontamination at room temperature and evaluate inactivation of a SARS-CoV-2 surrogate and a non-enveloped model virus as well as mask and respirator integrity following active multiple-cycle vaporised hydrogen peroxide (VHP), ultraviolet germicidal irradiation (UVGI), and dry heat (DH) decontamination.

**Methods:** Masks and respirators, inoculated with infectious porcine respiratory coronavirus or murine norovirus, were submitted to passive decontamination or single or multiple active decontamination cycles; viruses were recovered from sample materials and viral titres were measured via TCID_50_ assay. In parallel, filtration efficiency tests and breathability tests were performed according to EN standard 14683 and NIOSH regulations.

**Results and Discussion:** Infectious porcine respiratory coronavirus and murine norovirus remained detectable on masks and respirators up to five and seven days of passive decontamination. Single and multiple cycles of VHP-, UVGI-, and DH were shown to not adversely affect bacterial filtration efficiency of masks. Single- and multiple UVGI did not adversely affect respirator filtration efficiency, while VHP and DH induced a decrease in filtration efficiency after one or three decontamination cycles. Multiple cycles of VHP-, UVGI-, and DH slightly decreased airflow resistance of masks but did not adversely affect respirator breathability. VHP and UVGI efficiently inactivated both viruses after five, DH after three, decontamination cycles, permitting demonstration of a loss of infectivity by more than three orders of magnitude. This multi-disciplinal approach provides important information on how often a given PPE item may be safely reused.

## INTRODUCTION

As the severe acute respiratory syndrome coronavirus 2 (SARS-CoV-2) pandemic accelerates, the supply of personal protective equipment (PPE) remains under severe strain. In particular, the surging global demand for disposable surgical face masks (SMs) and filtering facepiece respirators (FFRs), identified as incremental for source control and prevention of onward transmission from infected individuals (SMs) and protection of health-care personnel during aerosol-generating procedures and support treatments (FFRs) (1–4), by far exceeds current manufacturing capacities.

To combat critical shortages, and in a departure from the prevailing culture of throwaway living (5) and a shift towards an eco-efficient circular economy within the healthcare industry (6), repeated re-use of typically single-use only items has been recommended (1,2,7,8). Prior decontamination is paramount to safe PPE re-use; SM and FFR reprocessing techniques must guarantee not only the complete inactivation of SARS-CoV-2 and other contaminating respiratory or oral human pathogens (the US Food and Drug Administration recommends a robust proof of infectious bioburden reduction of three orders of magnitude for viral pathogens (9)), but must do so without compromising the integrity of the items themselves.

In the context of a limited re-use strategy, CDC-issued reccommendations include storage of SMs or FFRs at room temperature (in a breathable paper bag) for a minimum period of five days of passive decontamination prior to re-use (10). However, SARS-CoV-2 room temperature survival rates have been subject to much debate, with earlier reports of an only short persistence (three or four days on porous and non-porous surfaces, respectively (11,12)) succeeded by more recent ones of significantly longer viability (21 days on PPE (13) and up to 28 days on various common surfaces (14)). While reported differences are likely dependent on multiple variables, including fluctuations in ambient temperature, relative humidity, light influx, and virus input, they certainly also reflect differences in the surfaces or carrier matrices themselves (15), necessitating targeted assays to evaluate and mitigate the individual risk of transmission via fomites in general and SMs or FFRs in particular.

Various studies have investigated active SM or FFR decontamination with regard to either biocidal efficacy (modelled utilising a wide range of organisms and matrices) (12,16) or the impact of repeat cycles on functional performance of SMs or FFRs (8,17–20). Few studies, however, offer a consolidated data set examining both viral inactivation as well as SM and FFR integrity subsequent to multiple-cycle decontamination (21). Current recommendations governing SM and FFR re-use are thus based on extrapolations from various sources describing assays performed under vastly differing experimental conditions and necessarily include not inconsiderable degrees of uncertainty (22–24).

Amongst the various SM or FFR reprocessing techniques under investigation, vaporised hydrogen peroxide (VHP), an industry standard chemical decontaminant implemented in medical-, pharmaceutical-, and research facilities, has garnered attention as a cost-effective and practical option for SM and FFR decontamination (8,9,17,21,22,25). Two physical decontamination methods, ultraviolet germicidal irradiation (UVGI) (18,19) and the application of dry heat (DH) (12,18), have further shown promise as SM or FFR reprocessing techniques.

We previously demonstrated efficient single-cycle VHP, UVGI, and DH decontamination of SMs and FFRs inoculated with two *in vitro* cultivable BSL2 pathogens. Inactivation of the infectious SARS-CoV-2 surrogate porcine respiratory coronavirus (PRCV) (26–30) demonstrated virucidal activity of all three methods against enveloped coronaviruses (31); decontamination of hardier non-enveloped human respiratory or oral pathogens, which can equally contaminate SMs or FFRs (9,32), was investigated using the notoriously tenacious murine norovirus model (MuNoV) (33–36).

Here we verify PRCV and MuNoV survivability rates on SMs and FFRs and investigate multiple-cycle active decontamination of coronavirus- or norovirus-inoculated SMs and FFRs, demonstrating that VHP, UVGI, and DH efficiently inactivate both viruses after several rounds of decontamination, all three methods inducing a loss of viral infectivity by more than three orders of magnitude in line with the FDA guidelines (9). In addition, an investigation into filtration efficiency and breathability of treated face coverings demonstrated that the cumulative use of UVGI, VHP, or DH did not adversely affect SM integrity following up to five decontamination cycles. Similarly, FFRs retained their integrity subsequent to five iterations of UVGI or VHP treatment; DH, however, was found to significantly alter the characteristics of FFRs when exceeding three decontamination rounds. Our multi-disciplinal, consolidated approach, wherein both virus inactivation and SM and FFR integrity are investigated subsequent to multiple decontamination cycles, provides important information on how often a given PPE item may be safely reused. This data provides a measure of security to health-care personnel and the general public; it can help close the currently existing gap between PPE supply and demand and can contribute to the development of circular economy policies in a post-Covid-19 era healthcare sector.

## MATERIALS AND METHODS

An overview of the workflow summarising the SM or FFR decontamination techniques, the number of applied cycles, and the tests to evaluate PPE integrity or virus inactivation, is provided in Figure 1.

**Figure 1.**
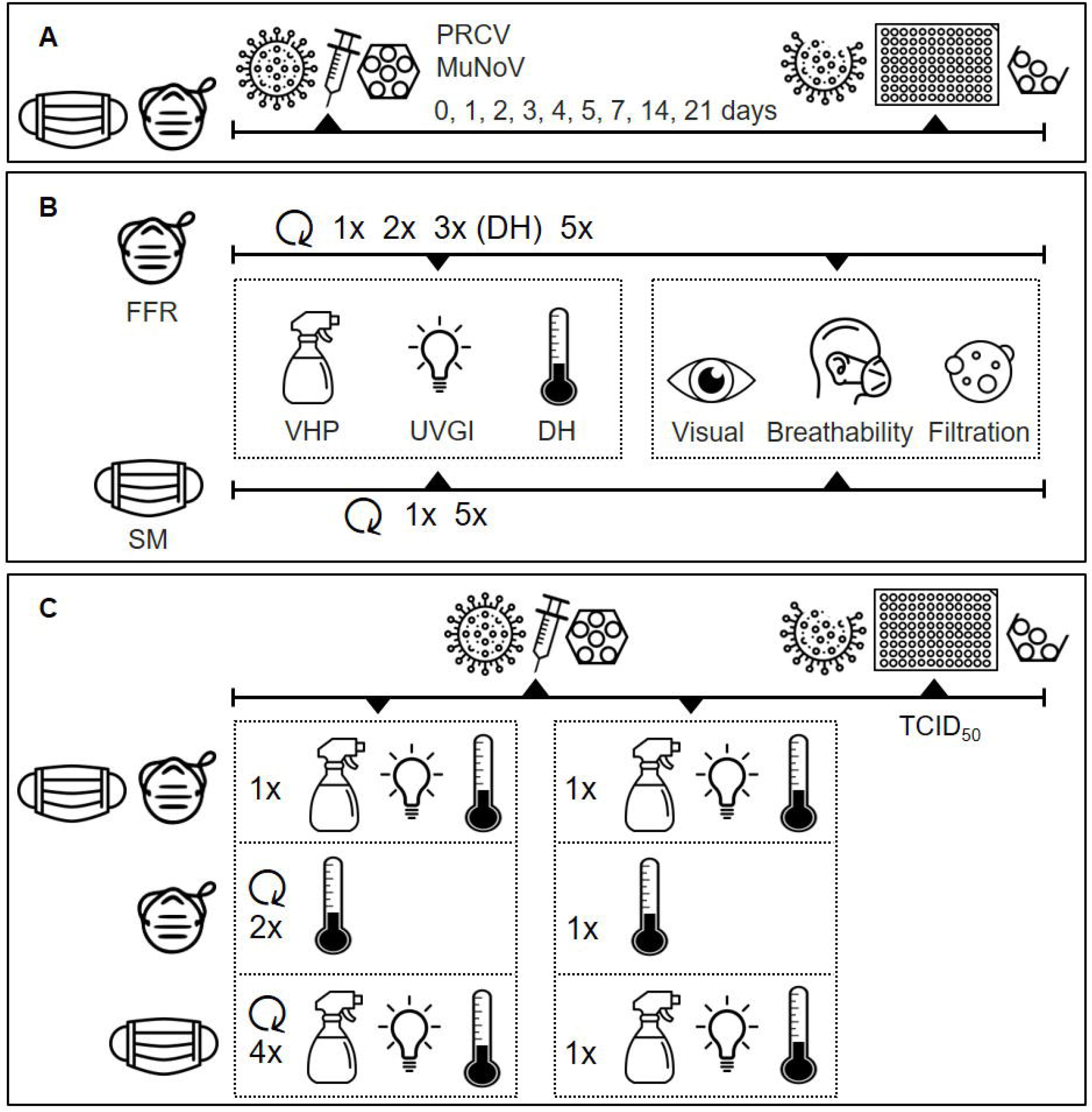
Experimental set-up of filtering facepiece respirator (FFR) and surgical mask (SM) decontamination assays. (A) Natural virus degradation over time. (B) Integrity testing after multiple-cycle vaporised hydrogen peroxide (VHP), ultraviolet germicidal irradiation (UVGI), and dry heat (DH) decontamination. (C) Multiple-cycle decontamination of porcine respiratory coronavirus (PRCV)- and murine norovirus (MuNoV)-inoculated SMs/FFRs.

### Surgical masks and filtering facepiece respirators

All FFRs and SMs were verified to be from the same respective manufacturing lot. Manufacturers (and models): KN95 FFR -Guangzhou Sunjoy Auto Supplies CO. LTD, Guangdong, China (2020 N°26202002240270); surgical mask (Type II) -Hangzhou Sunten Textile Co., LTD, Hangzhou, China (SuninCare™, Protect Plus).

### Decontamination techniques

#### Vaporised hydrogen peroxide

Surgical masks, FFRs and a chemical indicator were placed in individual Mylar/Tyvek pouches. Vaporous hydrogen peroxide treatment was performed with the V-PRO Max Sterilizer (Steris, Mentor, OH) which uses 59% liquid hydrogen peroxide to generate vapor. A 28-minute non lumen cycle consisted of 2 min 40 sec conditioning (5 g/min), 19 min 47 sec decontamination (2.2 g/min) and 7 min 46 sec aeration, with a peak VHP concentration of 750 ppm.

#### Ultraviolet germicidal irradiation

Surgical masks and FFRs were individually irradiated using a LS-AT-M1 (LASEA Company, Sart Tilman, Belgium) equipped with 4 UV-C lamps of 5.5W (@UV-C). Hung vertically on a metal frame, masks and FFRs were inserted into a safety enclosure. A 2 min UV-C treatment (surgical masks) led to a fluence of 2.6J/cm^2^ per mask (1.3J/cm^2^ per side). Power and irradiation time (120 s) were monitored and recorded throughout. Following irradiation, surgical masks and FFRs were unloaded and placed in individual bags.

#### Dry heat

Surgical masks and FFRs hung horizontally on a metal frame were inserted into an electrically heated vessel (M-Steryl, AMB Ecosteryl Company, Mons, Belgium) for 60 min (± 15 min) of heat treatment at 102°C (± 4°C) following the “Guidance for the reprocessing of SMs and FFRs during the coronavirus disease (COVID-19) Public Health Emergency” by the Belgian Federal Agency for Medicines and Health Products. Temperatures inside the heated vessel were recorded throughout to ensure correct exposure conditions. After termination of the treatment cycle, masks and FFRs were allowed to cool and then bagged individually.

### Surgical mask integrity testing

Integrity of decontaminated SMs was determined via initial macroscopic observation followed by EN 14683 standard filtration efficiency and breathability tests. Three SMs were used to analyse bacterial filtration efficiency (BFE), five to measure breathability.

#### SMs -Macroscopic observation

All SM performance testing was carried out at the Centexbel Textile Research Centre (Belgium). An initial visual inspection of SMs was carried out to verify their integrity; particular attention was paid to potential signs of degradation such as discoloration or deformation.

#### SMs -Bacterial filtration efficiency

BFE employs a ratio of upstream bacterial challenge to downstream residual concentration to determine filtration efficiency of SM materials against droplets. It is a required quantitative test method for SM clearance by the United States FDA and the European Medical Device Directive 93/42/EEC (BFE ≥ 98% according to EN 14683 for Type II and ASTM F2100 for Level 2 SMs). Briefly, SMs were conditioned at 85 ± 5 % relative humidity and 21 ± 5 °C prior to testing. BFE was measured using unneutralized *Staphylococcus aureus* bacteria contained within an aerosol droplet with a mean particle size of 3 µm diameter. The aerosol sample was drawn through an unfolded SM clamped to the top of a 6-stage Andersen impactor with agar plates for collection of the bacteria particles at a flow rate of 28.3 L/min for 1 min as per FDA guidance and ASTM F2101 method (challenge level of 1500 and 3000 colony-forming units (CFU) per test). Following removal and incubation of the culture plates, colonies were counted to determine total CFU and BFE. A positive control without a test filter sample clamped into the system was used to determine the number of viable particles used per test. A negative control with no bacteria in the airstream was performed to determine the background challenge in the glass aerosol chamber prior to testing.

#### SMs -Breathability

Breathability of SMs, defined as the measure of differential pressure required to draw air through a measured surface area at a constant air flow rate, was measured according to EN 14683 + AC:2019 (breathability < 40 Pa/cm^2^ for Type I and II; < 60 Pa/cm^2^ for Type IIR) (37). Briefly, a constant airflow of 8 L/min was applied through a 25 mm diameter holder (4.9 cm^2^ total surface area at orifice) to a SM test specimen. A mass flow controller was used to measure the flow rate and the the air exchange pressure of the SM material was measured using two manometers positioned upstream and downstream of the airflow. Measurements were performed on five SMs and five different locations per unfolded mask (top left, top right, bottom left, bottom right, and middle). The differential pressure per mask, expressed in Pa/cm^2^ and obtained by dividing pressure difference by surface area, was reported as the average of all twenty-five measurements (5 measurements per mask; 5 masks tested).

### Filtering facepiece respirator integrity testing

In the field of protective equipment, the nomenclature and standardisation pertaining to FFRs and their accreditation differ from one continent to another and even from one country to another. FFRs are generally referred to as FFP masks in Europe, KN95s in China, and N95s in the United States; the EN 149 + A1:2009 standard (primarily) and an ISO 16900 standard (to a lesser extent) are applied in Europe, National Institute for Occupational Safety and Health (NIOSH) procedures are invoked in the United States. While the different methods do not always have the same standardisation limits, the utilised techniques are generally the same. In the present study, FFR filtration efficiency and breathability tests were performed following NIOSH procedures. Three FFRs were used per test condition (assays performed in triplicate).

#### FFRs -Macroscopic observation

All FFR performance testing was carried out at the Nelson Laboratories (USA). An initial visual inspection of FFRs was carried out to verify their integrity; particular attention was paid to potential signs of degradation such as discoloration or deformation.

#### FFRs -NaCl filtration efficiency

FFR filtration efficiency was measured using the NIOSH sodium chloride (NaCl) aerosol method employed for certification of particulate respirators with an efficiency of ≥95% (42 CFR Part 84). Briefly, FFRs were pre-conditioned at 85 ± 5% relative humidity and 38 ± 2.5°C for 25 ± 1 hr prior to measurements. A NaCl solution was aerosolized (by atomising an aqueous solution of the salt and evaporating the water), charge neutralized, and then passed through the convex side of the FFRs. The concentrations of NaCl aerosol upstream and downstream of the FFR were measured at 85 L/min flow rate using a flame photometer, allowing for precise determinations in the range < 0.001 % to 100 % filter penetration.

#### FFRs -Breathability

FFR breathability was assessed using inhalation and exhalation breathing resistance measurements according to NIOSH 42CFR Part 84. Inhalation and exhalation resistance was tested according to NIOSH Standard Test Procedures (TEB-APR-STP-0007 and TEB-APR-STP-0003 (38)); results in mm H_2_O were recorded and evaluated against NIOSH performance criteria for FFR approvals (35 mm H_2_O for inhalation and 25 mm H_2_O for exhalation) at approximately 85 ± 2 L/min airflow.

### Virus inactivation testing

Virus infectivity losses at room temperature (passive decontamination) as well as the efficacy of VHP, UVGI, and DH in inactivating infectious PRCV or MuNoV after multiple SM or FFR decontamination cycles (active decontamination) were assessed using experimentally inoculated SMs and FFRs.

### Viruses and cells

The continuous swine testicle (ST) cell-line, grown from testicular foetal swine tissues as described by McClurkin and Norman (1966) (39), was maintained in MEM (GIBCO), supplemented with 5% foetal calf serum (FCS) (Sigma), 1% sodium pyruvate 100x (GIBCO), and antibiotics (100U/ml penicillin, 0.1mg/ml streptomycin and 0.05 mg/ml gentamycin).

PRCV strain 91V44 (40) was passaged three times on confluent ST monolayers. Titres were determined via the tissue culture infective dose (TCID_50_) method; ST cells were seeded in 96-well plates and infected with 10-fold serial dilutions of PRCV and incubated for four days at 37 °C with 5% CO_2_. Four days after inoculation, monolayers were analysed for the presence of cytopathic effect by light microscopy. Titres, expressed as TCID_50_/ml, were calculated according to the Reed and Muench transformation (41). PRCV stocks with a titre range of 2.00×10^7^ to 2.00×10^8^ TCID_50_/mL were used in subsequent steps.

The murine macrophage cell line RAW264.7 (ATCC TIB-71) was maintained in Dulbecco’s modified Eagle’s medium (Invitrogen) containing 10% FCS (BioWhittaker), 1% 1 M HEPES buffer (pH 7.6) (Invitrogen), and 2% of an association of penicillin (5000 SI units/ml) and streptomycin (5 mg/ml) (PS, Invitrogen) at 37 °C with 5% CO_2_.

Stocks of MuNoV isolate MNV-1.CW1 were produced by infection of RAW264.7 cells at a multiplicity of infection of 0.05. Two days post-infection, cells and supernatant were harvested and clarified by centrifugation for 10 minutes at 4000 × *g* after three freeze/thaw cycles (– 80°C/37°C). Titres were determined via the TCID_50_ method; RAW 264.7 cells were seeded in 96-well plates, infected with 10-fold serial dilutions of MuNoV, incubated for three days at 37 °C with 5% CO_2_, and finally stained with 0.2% crystal violet for 30 minutes. Titres, expressed as TCID_50_/ml, were calculated according to the Reed and Muench transformation (41). MuNoV stocks with a titre range of 2.00×10^6^ to 1.12×10^7^ TCID_50_/mL were subsequently used.

### Passive decontamination and multiple-cycle active decontamination of porcine respiratory coronavirus- or murine norovirus-inoculated surgical masks and filtering facepiece respirators

Assays investigating time-dependent effects of virus degradation at room temperature (passive decontamination), were performed using new SMs or FFRs. Per time point (0 hour, 1 day, 2 days, 3 days, 4 days, 5 days, 7 days, 14 days, and 21 days) and per virus (PRCV or MuNoV), one SM or FFR was inoculated. The workflow followed previously described protocols for SM and FFR inoculation and virus elution (31,33). Briefly, per SM or FFR, 100 µl of undiluted viral suspension were injected under the first outer layer at the centre of each of three square coupons (34 mm × 34 mm) previously outlined in graphite pencil on the intact SMs or FFRs. In addition to inoculation of the *de facto* SMs or FFRs, 100 µl of viral suspension were pipetted onto both elastic straps. SMs and FFRs thus inoculated were allowed to dry for 20 minutes at room temperature in a class II biological safety cabinet and were then incubated in the dark (to limit any effect light might have on viral decay) at laboratory room temperature (average 20°C) for the specified time points.

Assays investigating cumulative effects of multiple-cycle VHP and UVGI on SM or FFR decontamination (active decontamination), consisted of either one or four decontamination cycles applied prior to PRCV or MuNoV inoculation and subsequent decontamination, thus resulting in an overall total of two and five decontaminations per SM or FFR. Since cumulative DH treatments were found to significantly alter the characteristics of FFRs when exceeding three decontamination cycles (see below), assays investigating cumulative effects of multiple-cycle DH decontamination, consisted of either one or two FFR decontamination cycles applied prior to PRCV or MuNoV inoculation and subsequent decontamination, resulting in a maximum number of three DH decontaminations. Per decontamination method and type of face covering within the respective assays, one negative control SM or FFR (uncontaminated but treated), three treated SMs or FFRs (PRCV- or MuNoV-contaminated and treated), and three positive controls (PRCV- or MuNoV-contaminated but untreated) were utilised. Per treated or control SM or FFR, 100 µl of undiluted viral suspension were injected under the first outer layer at the centre of each of three square coupons. In addition to inoculation of the *de facto* SMs or FFRs, 100 µl of viral suspension were pipetted onto one elastic strap per contaminated SM or FFR. SMs and FFRs were allowed to dry for 20 minutes at room temperature in a class II biological safety cabinet before final decontamination via UVGI, VHP, or DH.

Upon completion of the different decontamination protocols, PRCV or MuNoV was eluted from three excised coupons and one severed elastic strap per SM or FFR (in the case of passive decontamination assays both straps) via maximum speed vortex (2500 revolutions per minute in a VWR VX-2500 Multi-Tube Vortexer; 1 minute- or 20 minute vortex for PRCV- and MuNoV inoculated SMs or FFRs, respectively) into 4 mL elution medium consisting of MEM or DMEM (Sigma)) supplemented with 2

% of an association of penicillin (5000 SI units/mL) and streptomycin (5 mg/mL) (PS, Sigma); for elution from VHP-treated SMs or FFRs, 20% FCS and 0.1% β-mercaptoethanol were added to the medium. Titres of infectious PRCV or MuNoV recovered from individual coupons and straps were determined via TCID_50_ assay. Back titrations of inoculum stocks were performed in parallel to each series of decontamination experiments.

### Data analysis and statistics

Statistical analyses of differences in infectious viral titres were performed using GraphPad Prism 7 (Graph-Pad Software) and P-values were computed by using a two-sided independent sample t-test, where ****P<0.0001, ***P<0.001, **P<0.01, *P<0.05, and ns is P≥0.05.

## RESULTS AND DISCUSSION

### Infectious porcine respiratory coronavirus is recovered up to five and seven days after inoculation of SMs and FFRs; murine norovirus remains detectable after seven days of passive SM or FFR decontamination

To combat PPE shortages provoked by the SARS-CoV-2 pandemic, repeated re-use of both SMs and FFRs has been recommended (1,2,7,8). Prior decontamination of SARS-CoV-2 and other respiratory or oral human pathogens is paramount to SM or FFR safe re-use and may be achieved either passively via storage of items or via active SM and FFR reprocessing.

To validate CDC-issued limited re-use recommendations for passive decontamination by storage (10), we evaluated time-dependent persistence of PRCV, an infectious SARS-CoV-2 surrogate, and MuNoV, a notoriously tenacious small non-enveloped oral pathogen, on SMs and FFRs. Infectious PRCV was detectable for up to five days post inoculation on SM coupons (1.52 (±0.38) log_10_ TCID_50_/mL) and three days post inoculation on SM straps (0.88 (±0.11) log_10_ TCID_50_/mL). The recovery of PRCV from FFRs was similar to that of SMs, with coupon virus levels near the assay LOD between days three and five post inoculation and 1.04 (±0.42) log_10_ TCID_50_/mL detected at day seven post inoculation; no infectious PRCV was recovered from straps past day one post inoculation (Figure 2). Infectious MuNoV remained detectable after seven days of passive SM or FFR coupon decontamination (1.88 (±0.38) and 0.97 (±0.14) log_10_ TCID_50_/mL, respectively) and was also elutable from SM and FFR straps at this time (1.43 (±0.53) and 1.18 (±0.18) log_10_ TCID_50_/mL, respectively) (Figure 3).

**Figure 2.**
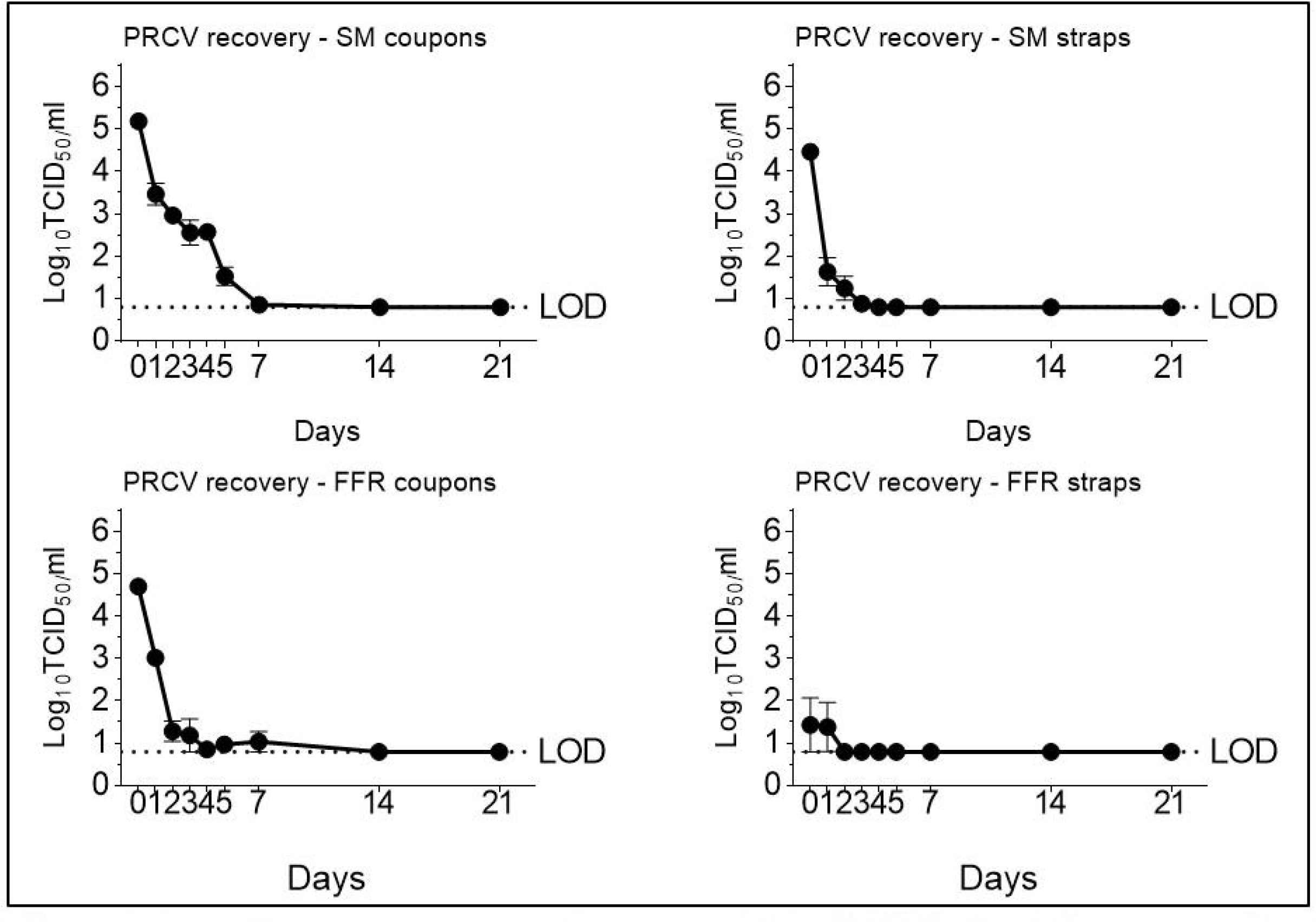
Recovery of porcine respiratory coronavirus (PRCV) after elution from filtering facepiece respirators (FFRs) and surgical masks (SMs) kept at room temperature (20°C) over time. PRCV infectivity was analysed in swine testicular cells. The cell culture limit of detection (LOD) was 0.80 log10 TCID_50_/mL (6.31×10^0^ TCID_50_/mL).

**Figure 3.**
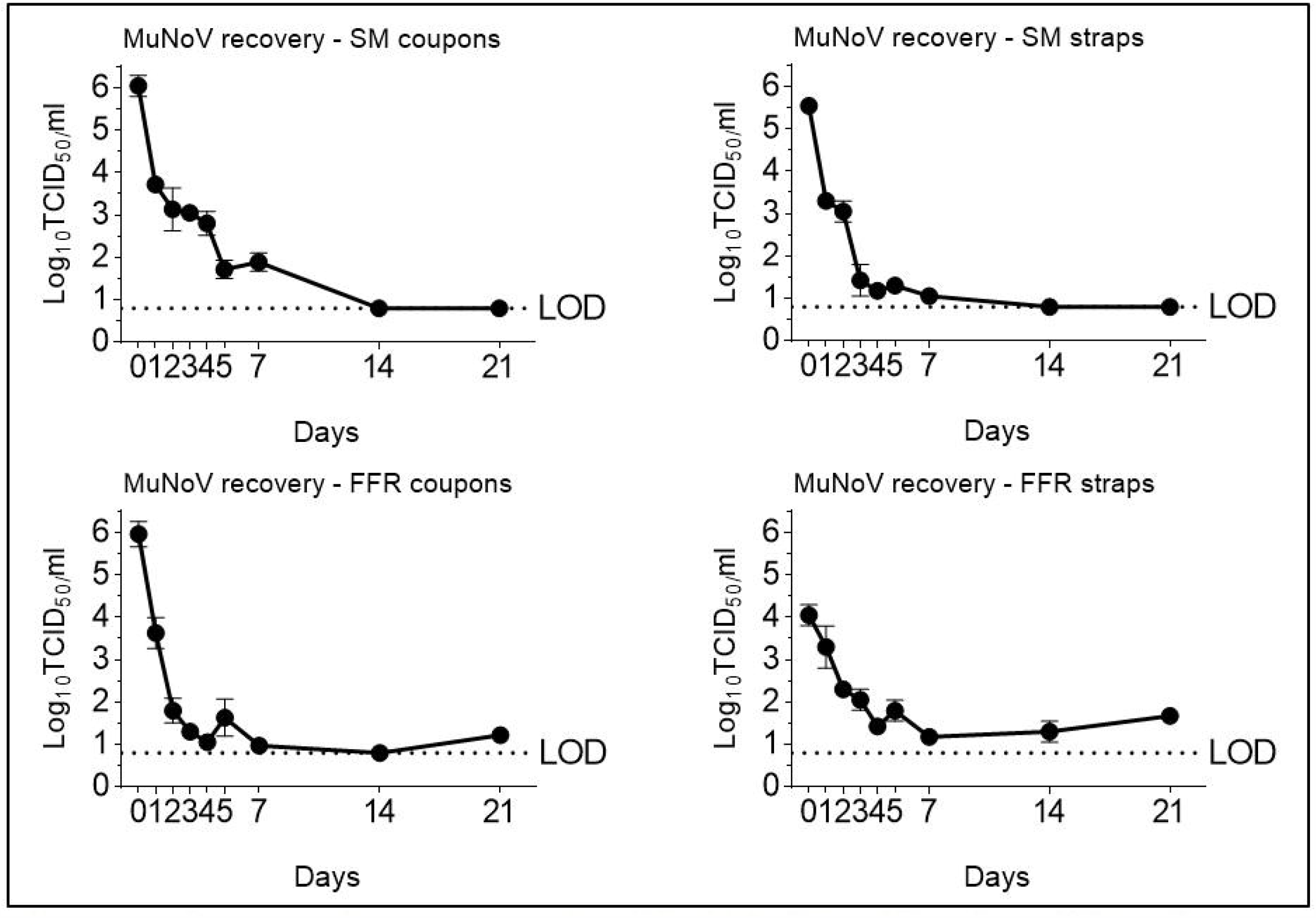
Recovery of murine norovirus (MuNoV) after elution from filtering facepiece respirators (FFRs) and surgical masks (SMs) kept at room temperature (20°C) over time. MuNoV infectivity was analysed in RAW264.7 cells. The cell culture limit of detection (LOD) was 0.80 log10 TCID_50_/mL (6.31×10^0^ TCID_50_/mL).

We confirm passive room temperature SM and FFR decontamination to be effective for both PRCV and MuNoV inactivation. However, we show that CDC-issued recommendations of a five-day room temperature storage (10) may be too short as they do not allow for total degradation of high virus loads on all SM and FFR materials (this in line with recent observations on other PPE items (13,14)). According to our observations, the storage period should ideally be extended to at least seven days for safe coronavirus inactivation and to a minimum of 14 days for decontamination of non-enveloped viruses such as noroviruses.

### Up to five cycles of active VHP and UVGI decontamination do not visually affect SMs or FFRs; up to five and up to three DH cycles do not affect the physical appearance of SMs and FFRs, respectively

In high-throughput environments that necessitate a ready PPE availability (hospitals, nursing homes, and other public facilities), an extended storage and turnaround time of one or even two weeks may not be feasible, necessitating the implementation of fast-acting active decontamination techniques.

Active decontamination must guarantee not only the inactivation of SARS-CoV-2 and other pathogens, but must do so without compromising the integrity of the SMs or FFRs themselves. Decontaminating treatments are known to have inherently detrimental side effects that may compromise the integrity of decontaminated objects (42); while VHP, UVGI, and DH decontamination have previously been shown to not significantly impact performance of polypropylene-based SMs or FFRs following single cycle decontamination (17–19,21), the maximum number of decontamination cycles may be limited (42). To validate repeated safe reuse of SMs and FFRs, we investigated SM integrity subsequent to one and five, and FFR integrity subsequent to one, two, and five cycles of VHP, UVGI, and DH decontamination.

After one VHP, UVGI or DH decontamination cycle, no abnormalities were registered at visual SM or FFR inspection. After multiple decontamination cycles VHP- or UVGI-treated SMs and FFRs remained physically unaffected, this in line with previous studies (43,44). Only FFRs subjected to five cycles of DH showed signs of degradation or burning which manifested as brown discoloration of FFR elastic straps and disassociation of the metal noseband from FFR fabrics; as a consequence, five cycles of DH treatment were abandoned in further analyses and were, uniquely for DH, replaced by tests performed after three treatment cycles.

### Single and multiple cycles of VHP-, UVGI-, and DH decontamination do not adversely affect SM BFE. Single- and multiple UVGI decontamination does not adversely affect FFR NaCl filtration efficiency, while VHP and DH treatments induce a slight decrease in filtration efficiency after one or three decontamination cycles

To investigate whether one and five and one, two, and five (three for DH) cycles of decontamination affect SM and FFR integrity, respectively, SM BFE testing was performed according to EN14683 and FFR filtration efficiency was investigated using the sub-micron NaCl aerosol method (NIOSH 42 CFR Part 84). Both SMs and FFRs surpassed minimum filtration efficiency requirements before (99.50% (±0.08) BFE and 97.01% (±0.56) NaCl filtration efficiency) decontamination. SM BFE remained consistently higher than 98% after single- and multiple-cycle decontamination (Figure 4 A).

**Figure 4.**
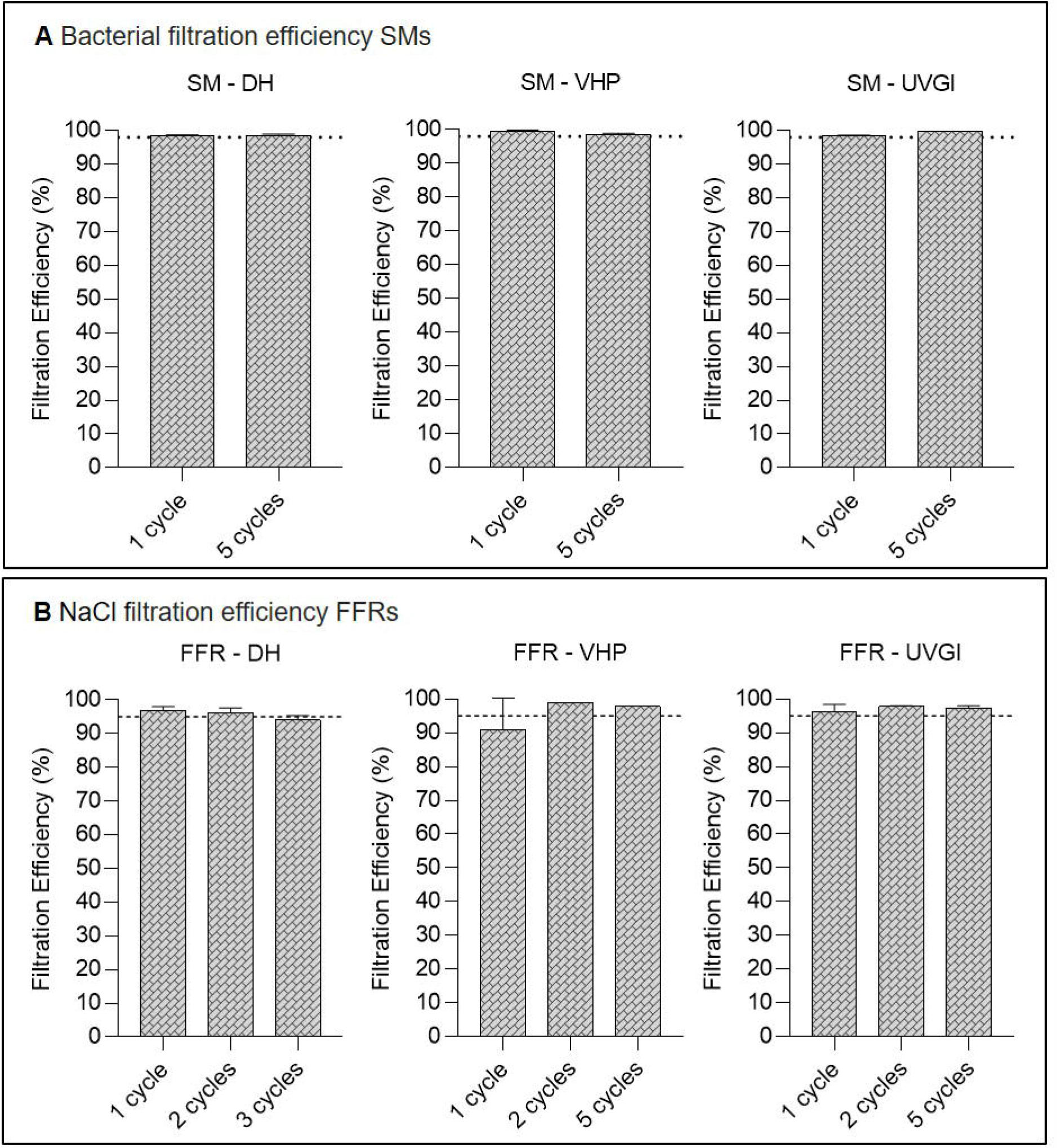
Filtering facepiece respirator (FFR) NaCl filtration efficiency- and surgical mask (SM) bacterial filtration efficiency (BFE) testing after single-cycle or multiple-cycle decontamination using dry heat (DH), vaporised hydrogen peroxide (VHP), and ultraviolet germicidal irradiation (UVGI). Horizontal dashed lines represent the NaCl filtration efficiency requirement of ≥95% according to NIOSH 42 CFR Part 84. Untreated FFRs (n=3) surpassed the minimum NaCl filtration efficiency, achieving 97.01% (±0.56) as a baseline before treatment. Horizontal dotted lines represent the bacterial filtration efficiency (3 µm droplet size) requirement of ≥98% according to EN 14683 for Type II and ASTM F2100 for Level 2 SMs. Untreated SMs (n=3) surpassed the minimum BFE, achieving 99.50% (±0.08) as a baseline before treatment.

FFR filtration efficiencies remained above the required ≥95% (i.e. <5% penetration) following DH and UVGI single-cycle treatments, however dropped to 91.02% (±8.38) post VHP exposure (this owing to the aberrant value of 79.2% for a single FFR). Following two, three (for DH), or five decontamination cycles, filtration efficiency of UVGI- and VHP-treated FFRs remained above 95%, but dropped to 94.16% (±1.02) after three cycles of DH decontamination (Figure 4 B). VHP (which is FDA-authorised for FFR decontamination) is typically not destructive to polypropylene FFRs (8,22) and has previously been shown to not negatively affect FFR performance after single or multiple decontamination cycles in assays similar to ours (43,45). Since neither two nor five cycles of decontamination caused a drop in filtration efficiency, it seems likely that the single aberrant result after one VHP cycle may have been due to an issue with the item itself rather than the decontamination. It follows that all three methods are suitable for single-cycle FFR decontamination and reuse and that UVGI- and VHP decontamination may safely be applied to FFRs for up to five cycles. DH at 102°C should only be used for a maximum of three iterations; for more than three DH decontamination cycles, only temperatures that preserve the filtration characteristics of pristine FFRs (< 100°C) are to be recommended (18,45).

### Multiple cycles of VHP-, UVGI-, and DH decontamination decrease airflow resistance of SMs but do not adversely affect FFR breathability

Breathability, or resistance to airflow during inhalation and exhalation, is an indication of the difficulty in breathing through SMs or FFRs and as such is important to wearer comfort. Breathability of SMs was measured via differential pressure (pressure drop) test according to EN 14683 + AC:2019 (37), while breathability of FFRs was assessed by inhalation and exhalation resistance tests according to NIOSH Standard Test Procedures (TEB-APR-STP-0007 and TEB-APR-STP-0003).

Untreated SMs (n=5) reached 52.08 (±0.99) Pa/cm^2^ differential pressure before treatment, while differential pressures were only slightly elevated following single-cycle DH (54.88 (±3.00) Pa/cm^2^) and VHP (59.2 (±3.88) Pa/cm^2^) decontamination, but exceeded the limit of 60 Pa/cm^2^ post UVGI treatment with a measurement of 63.72 (±7.05) Pa/cm^2^ (Figure 5). Following five decontamination cycles, pressure drop test results consistently exceeded the prescribed maximum of 60 Pa/cm^2^ (Figure 5), with mean values of 66.82 (±2.88) Pa/cm^2^ (DH), 69.04 (±3.88) Pa/cm^2^ (VHP) and 59.78 (±1.47) Pa/cm^2^ (UVGI). Such elevated results should exclude the tested SMs from use following multiple-cycle decontamination via all three methods according to EN 14683 + AC:2019; however, it should be noted that mean differential pressure results have been shown to vary depending on the SM type analysed (45). Hence, values exceeding the 60 Pa/cm^2^ limit in this study may have been artificially elevated by high SM baseline values prior to decontamination rather than the decontamination proceedures themselves, which have, in other studies, been shown to retain high SM performance even after multiple treatment cycles (45,46). In Belgium, where SMs may be marketed and used in the Covid-19 crisis situation according to an “Alternative Test Protocol” issued by the Belgian Federal Agency for Medicines and Health Products that sets the maximum differential pressure limit at ≤ 70 Pa/cm^2^ (47), all treated SMs met current breathability requirements. FFR inhalation and exhalation resistance measurements remained far below the recommended maximum limits of ≤35 mmH_2_O in inhalation and ≤25 mmH_2_O in exhalation maintaining acceptable respirability according to applicable standards and regulations both before (inhalation: 12.43 (±0.69) mmH_2_O; exhalation: 11.9 (±0.86) mmH_2_O) and after single or multiple decontamination cycles, echoing other published results (45,48).

**Figure 5.**
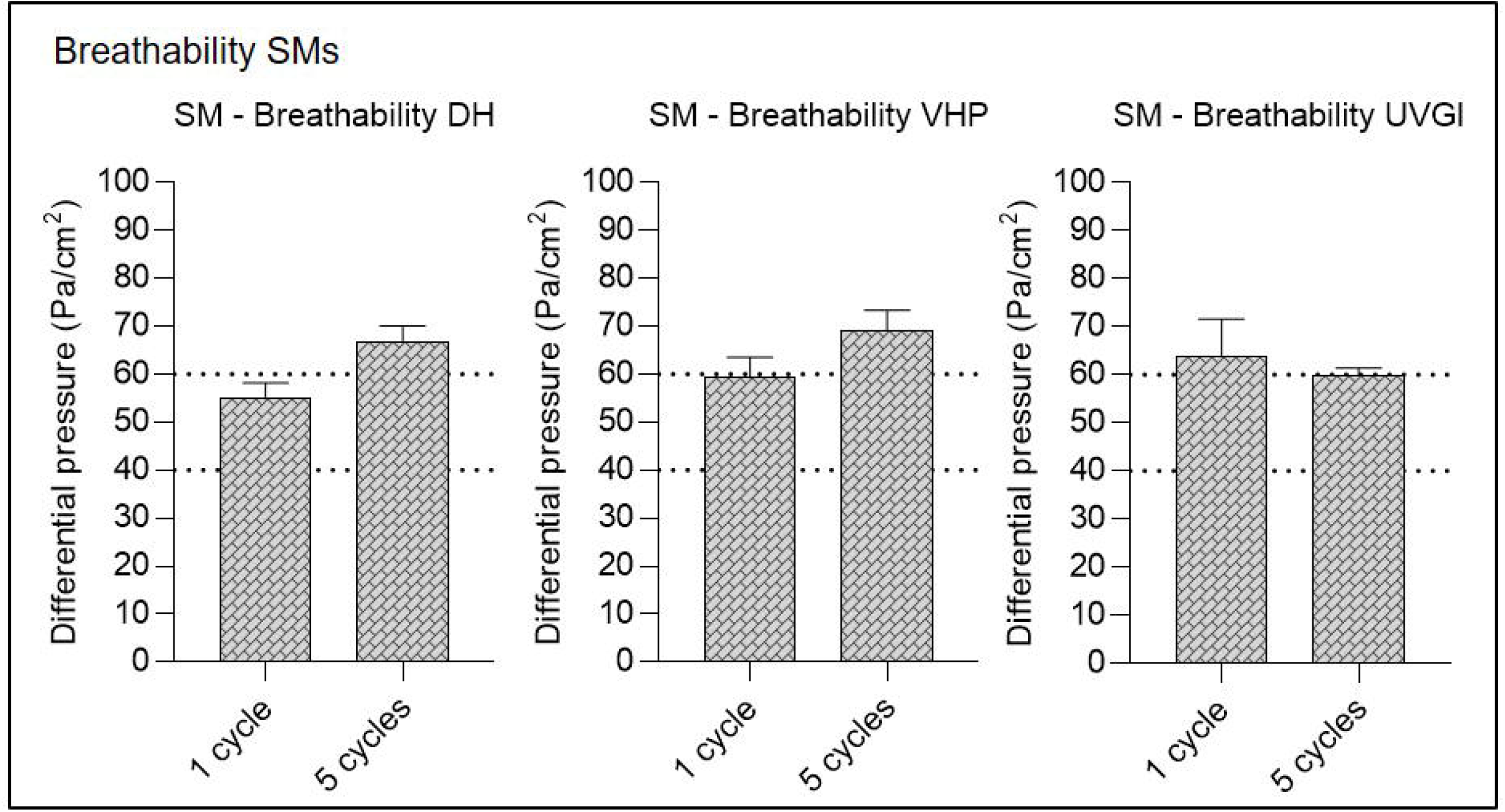
Surgical mask (SM) breathability testing after single-cycle or multiple-cycle decontamination using dry heat (DH), vaporised hydrogen peroxide (VHP), and ultraviolet germicidal irradiation (UVGI). Horizontal dotted lines represent the maximum allowed differential pressure in following standards: <40 Pa/cm^2^ according to EN 14683:2019 Annex C for Type I and II masks and < 60 Pa/cm^2^ for Type IIR. Untreated SMs (n=5) achieved 52.08 (±0.99) Pa/cm^2^ differential pressure as a baseline before treatment.

### Infectious porcine respiratory coronavirus is recovered at high titres from positive control SM- and FFR coupons, at lower titres from straps, and remains under the limit of detection following two (VHP, UVGI, DH), three (DH-treated FFRs) or five (VHP, UVGI, DH (SM)) active decontamination cycles

#### PRCV recovery from SM and FFR positive controls

Back titrations of virus inoculums performed in parallel to each series of experiments confirmed PRCV inoculum titres to be within a range of 7.30 to 8.30 log_10_ TCID_50_/mL for all experiments. The cell culture limit of detection (LOD) was 0.8 log_10_ TCID_50_/mL for all assays. An initially observed VHP cytotoxicity and correspondingly elevated LOD of 1.80 log_10_ TCID_50_/mL of VHP-treated coupon eluates was corrected via β-mercaptoethanol and FCS supplementation of elution medium; elevated cytotoxicity of VHP-treated strap eluates (SM and FFR) could not be neutralised and remained at 1.80 log_10_ TCID_50_/mL. Values below the LOD were thus considered as ≤0.80 log_10_ TCID_50_/mL or ≤1.80 log_10_ TCID_50_/mL (VH-treated straps). Comparable high levels of infectious virus were recovered from once-, twice-(DH-treated FFRs) or four-times treated, PRCV-inoculated left, right and middle coupons of all SMs and FFRs within a range of 4.27 (±0.50) to 6.07 (±0.29) log_10_ TCID_50_/mL (Supplementary Figure 1). Recovery values for infectious PCRV from SM and FFR straps were also similar between experiments, however they were lower than coupon recovery values, with mean values ranging from below the LOD to 4.44 (±0.74) log_10_ TCID_50_/mL (Supplementary Figure 1).

#### Multiple cycle decontamination of PRCV-inoculated SMs

Following two cycles of SM UVGI, VHP exposure, and DH treatment, all PRCV titres remained below the respective LOD of the assay (with the exception of UVGI treated straps), showing a total loss of infectivity of more than five orders of magnitude for UVGI-treated coupons (5.05 log_10_ reduction) and four orders of magnitude for VHP- and DH-treated coupons (4.83 and 4.39 log_10_ reduction, respectively), this in line with previous publications (48,49). Titres of PRCV recovered from SM straps following two treatment cycles were reduced by over two orders of magnitude post UVGI, VHP and DH treatment of SM straps (2.48, 2.22 and 2.85 log_10_ reduction) (Figure 7).

**Figure 6.**
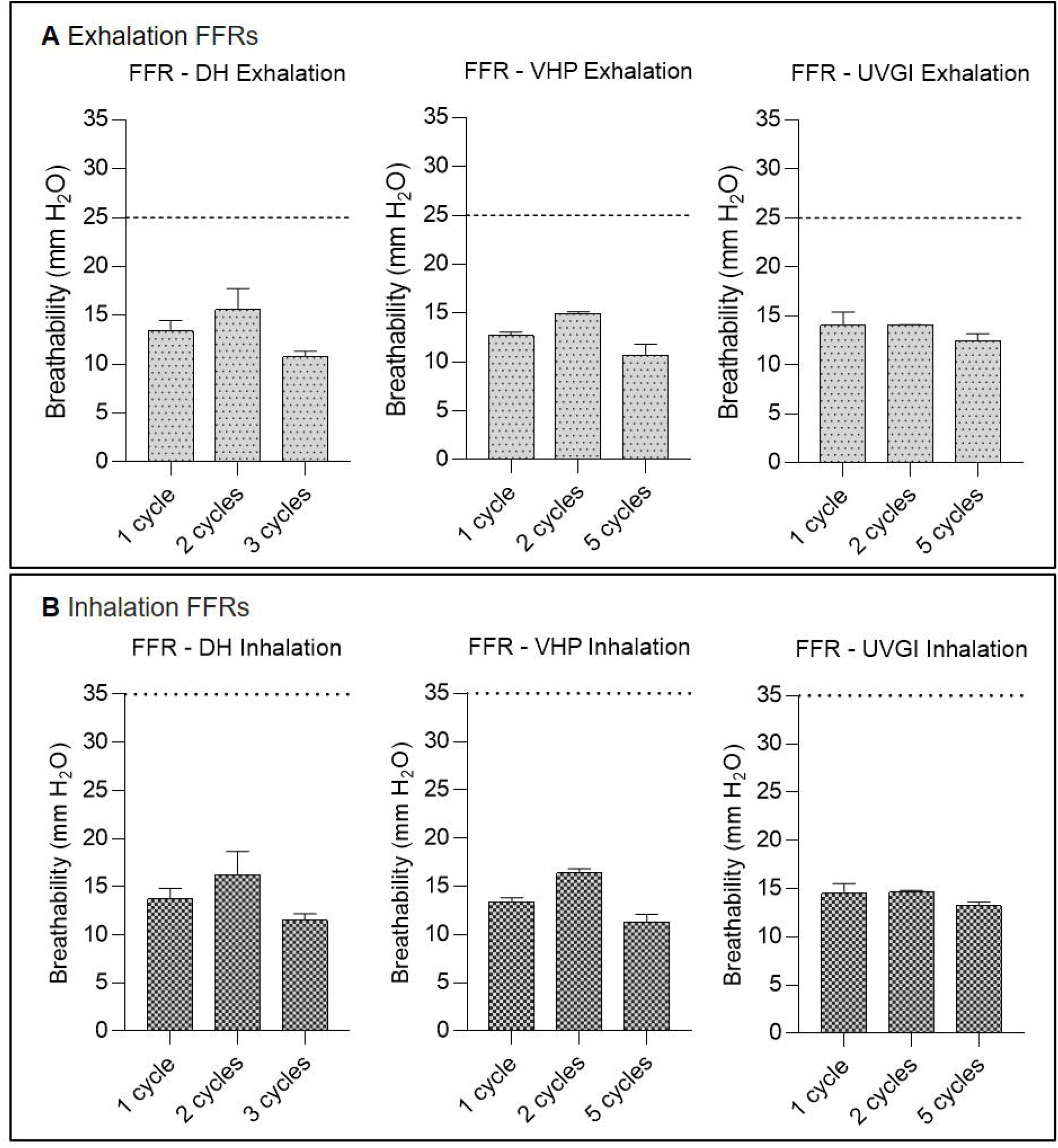
Filtering facepiece respirator (FFR) breathability testing after single-cycle or multiple-cycle decontamination using dry heat (DH), vaporised hydrogen peroxide (VHP), and ultraviolet germicidal irradiation (UVGI). Exhalation (A) and inhalation (B) breathing resistances after decontamination. Horizontal dashed (above) and dotted (below) lines represent the following breathing resistance standards: Exhalation: ≤25 mmH_2_O and Inhalation: ≤35 mmH_2_O for FFRs according to NIOSH 42 CFR Part 84. Untreated FFRs (n=5) achieved inhalation and exhalation resistance of 12.43 (±0.69) mmH_2_O and 11.9 (±0.86) mmH_2_O, respectively.

**Figure 7.**
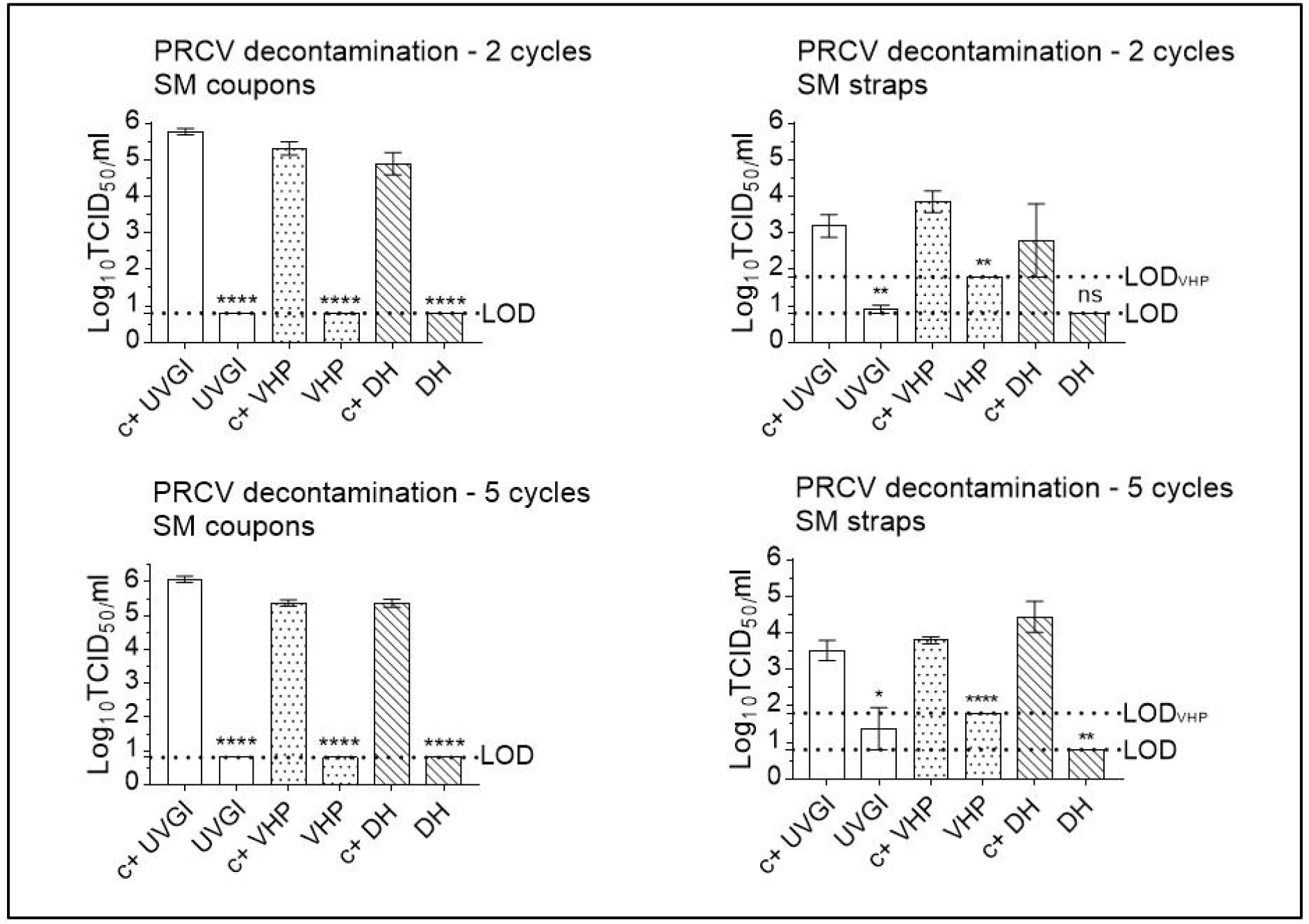
Porcine coronavirus (PRCV) inactivation following multiple cycle surgical mask (SM) decontamination using dry heat (DH), vaporised hydrogen peroxide (VHP), and ultraviolet germicidal irradiation (UVGI). Titrations were performed after two or five (three in the case of DH) decontamination treatments on PRCV-inoculated SM coupons and straps. PRCV infectivity was analysed in swine testicular cells. The cell culture limit of detection (LOD) was 0.80 log10 TCID_50_/mL (6.31×10^0^ TCID_50_/mL) for all analyses except those concerning VHP-treated SM straps (1.80 log10 TCID_50_/mL (6.31×10^1^ TCID_50_/mL)). Per decontamination method, nine PRCV-inoculated, decontaminated coupons (n=9) and three inoculated, decontaminated straps (n=3) were analysed in parallel to inoculated, untreated, positive control coupons (n=9) and straps (n=3). Mean log10 TCID_50_/mL and standard errors of the means are represented. P-values were computed by using a two-sided independent sample t-test, where ****P<0.0001, ***P<0.001, **P<0.01, *P<0.05, and ns is P≥0.05.

Following five cycles of SM UVGI, VHP exposure, and DH treatment, all PRCV titres remained below the respective LOD of the assay (with the exception of UVGI treated straps), showing a total loss of infectivity of more than five orders of magnitude for UVGI-treated coupons (5.37 log_10_ reduction) and more than four orders of magnitude for VHP- and DH-treated coupons (4.64 and 4.69 log_10_ reduction, respectively); titres of PRCV recovered from treated SM straps were reduced by over one order of magnitude post UVGI (1.59 log_10_ reduction) and for VHP-treated straps (2.02 log_10_ reduction), and by almost four orders of magnitude for DH-treated straps (3.94 log_10_ reduction) (Figure 7).

#### Multiple cycle decontamination of PRCV-inoculated FFRs

Decontamination treatment effects followed a similar pattern of PRCV inactivation for FFR coupons decontaminated twice via DH, VHP, and UVGI reducing viral titres by more than three and four orders of magnitude (3.71, 4.45 and 4.62 log_10_ reduction, respectively), supporting previous observations (31). The impact of two-cycle decontamination could not be measured for DH-treated FFR straps due to insufficient recovery of infectious virus in the corresponding controls. Virus recovery from both SM and FFR straps has been shown to be highly variable both in our hands (31) and in those of others (50) (and indeed, probably for this reason, strap decontamination is rarely assessed). Without enough proof of inactivation, we cannot recommend safe decontamination of SM or FFR straps and suggest treating straps separately using a disinfecting wipe or similar approach. Two-cycle UVGI and VHP treatment of FFR straps resulted in a reduction of infectious PRCV loads by 1.46 and 0.63 log_10_ reduction, respectively (Figure 8).

**Figure 8.**
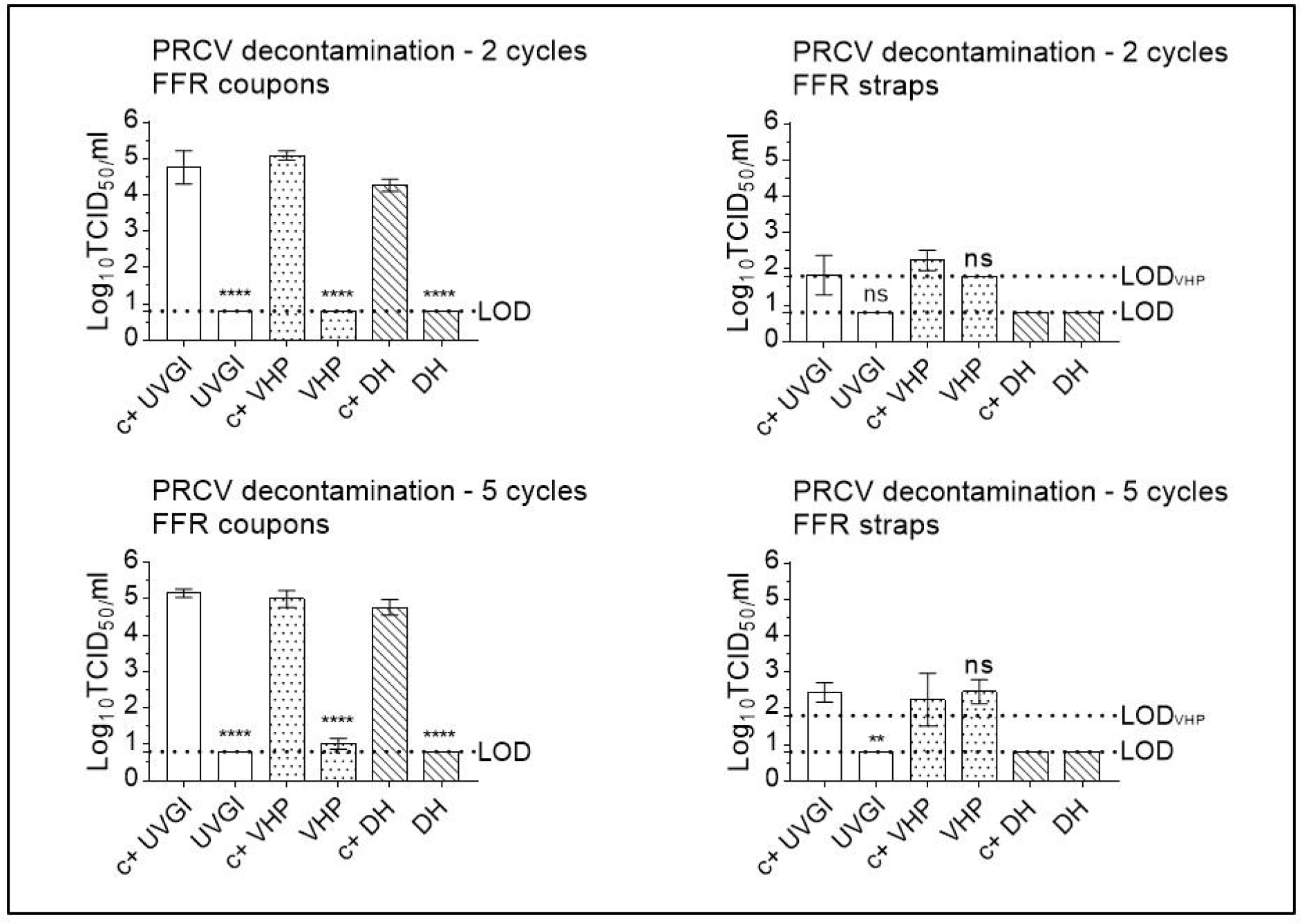
Porcine coronavirus (PRCV) inactivation following multiple cycle filtering facepiece respirator (FFR) decontamination using dry heat (DH), vaporised hydrogen peroxide (VHP), and ultraviolet germicidal irradiation (UVGI). Titrations were performed after two or five (three in the case of DH) decontamination treatments on PRCV-inoculated FFR coupons and straps. PRCV infectivity was analysed in swine testicular cells. The cell culture limit of detection (LOD) was 0.80 log10 TCID_50_/mL (6.31×10^0^ TCID_50_/mL) for all analyses except those concerning VHP-treated FFR straps (1.80 log10 TCID_50_/mL (6.31×10^1^ TCID_50_/mL)). Per decontamination method, nine PRCV-inoculated, decontaminated coupons (n=9) and three inoculated, decontaminated straps (n=3) were analysed in parallel to inoculated, untreated, positive control coupons (n=9) and straps (n=3). Mean log10 TCID_50_/mL and standard errors of the means are represented. P-values were computed by using a two-sided independent sample t-test, where ****P<0.0001, ***P<0.001, **P<0.01, *P<0.05, and ns is P≥0.05.

Following five cycles of FFR UVGI, VHP, and DH, all PRCV titres remained below the respective LOD of the assay, reducing viral titres by over four orders of magnitude (4.48, 4.22 and 4.30 log_10_ reduction, respectively). These results are in line with our own and others’ prior publications regarding decontamination of SARS-CoV-2- or surrogate-contaminated FFRs (31,49) and confirm that all three methods yield rapid and efficient virus inactivation even after multiple-cycle FFR decontamination. The impact of decontamination could not be measured for DH-treated FFR straps due to insufficient recovery of infectious virus in the corresponding controls. UVGI and VHP treatment of FFR straps resulted in a reduction of infectious PRCV loads by 1.81 and 0.18 log_10_ reduction, respectively (Figure 8).

### Infectious murine norovirus is recovered at high titres from positive control SM- and FFR coupons, at lower titres from straps, and remains under the limit of detection following two (VHP, UVGI, DH), three (DH) or five (VHP, UVGI) decontamination cycles

#### MuNoV recovery from SM and FFR positive controls

Back titrations of virus inoculums performed in parallel to each series of experiments confirmed MuNoV inoculum titres to be within a range of 6.30 to 7.05 log_10_ TCID_50_/mL for all experiments. The cell culture limit of detection (LOD) was 0.80 log_10_ TCID_50_/mL for all assays except for those concerning VHP-treated SM- or FFR straps and UVGI-treated FFR straps (1.80 log_10_ TCID_50_/mL). Comparable high levels of infectious virus were recovered from once-, twice-(DH-treated FFRs) or four-times treated, MuNoV-inoculated left, right and middle coupons of all SMs and FFRs within a range of 4.55 (±0.60) to 5.38 (±0.25) log_10_ TCID_50_/mL (Supplementary Figure 2). Recovery values for infectious MuNoV from SM and FFR straps were also similar between experiments, however they were lower than coupon recovery values, with mean values ranging from 1.80 (VHP LOD) to 5.22 (±0.14) log_10_ TCID_50_/mL (Supplementary Figure 2).

#### Multiple cycle decontamination of MuNoV-inoculated SMs

Following two cycles of SM UVGI, VHP exposure, and DH treatment, all MuNoV titres remained below the respective LOD of the assay, showing total loss of infectivity of over four orders of magnitude for UVGI-, VHP- and DH-treated SM coupons (4.47, 4.33, and 4.15 log_10_ reduction, respectively). Titres of MuNoV recovered from treated SM straps were reduced by less than three orders of magnitude post two cycles of UVGI and VHP treatment (0.96 and 2.55 (below the LOD) log_10_ reduction, respectively) and by over four orders of magnitude post two-cycle-DH treatment (4.43 log_10_ reduction (below LOD)) (Figure 9).

**Figure 9.**
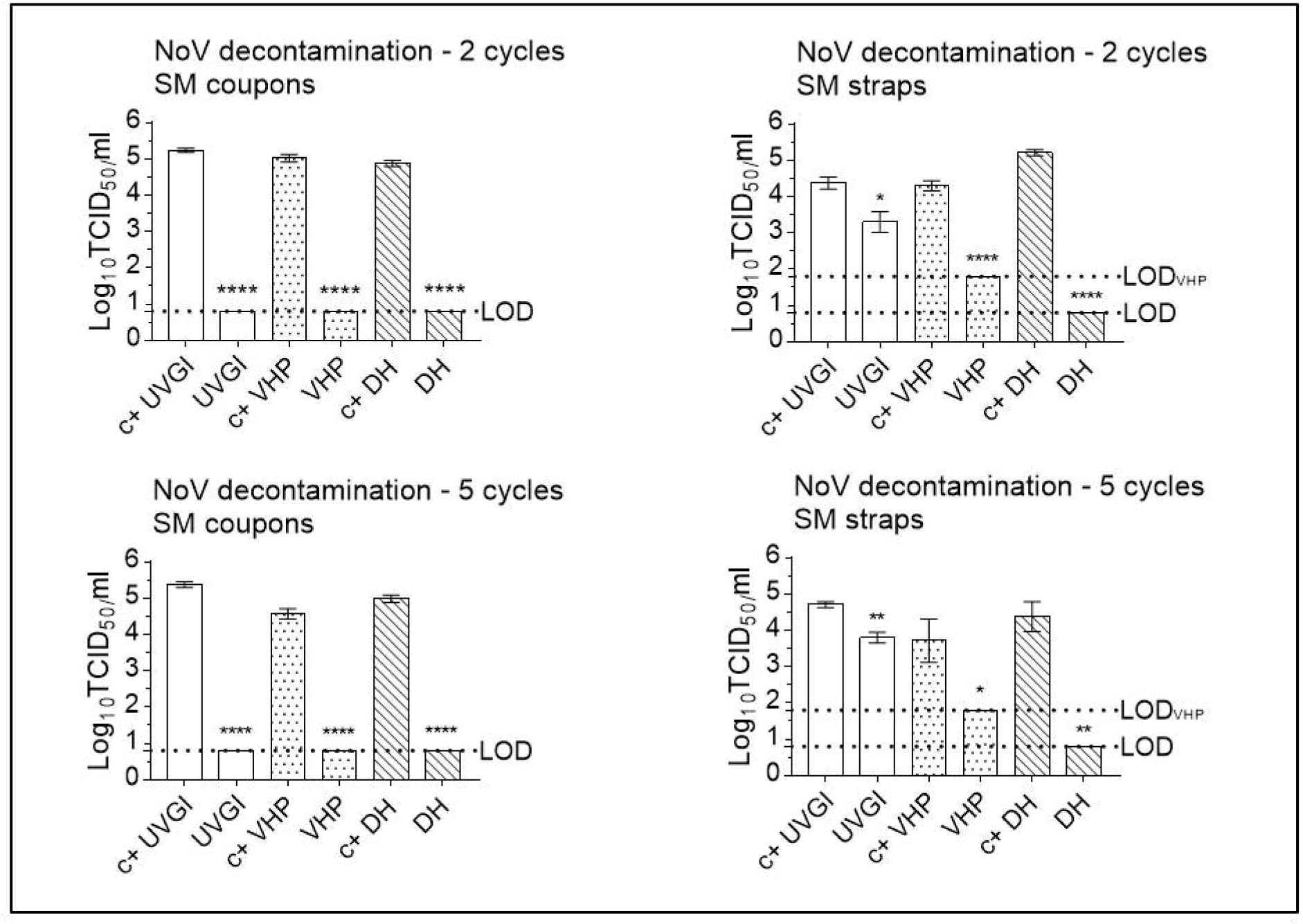
Murine norovirus (MuNoV) inactivation following multiple cycle surgical mask (SM) decontamination using dry heat (DH), vaporised hydrogen peroxide (VHP), and ultraviolet germicidal irradiation (UVGI). Titrations were performed after two or five (three in the case of DH) decontamination treatments on MuNoV-inoculated SM coupons and straps. MuNoV infectivity was analysed in RAW264.7 cells. The cell culture limit of detection (LOD) was 0.80 log10 TCID_50_/mL (6.31×10^0^ TCID_50_/mL) for all analyses except those concerning VHP-treated SM straps (1.80 log10 TCID_50_/mL (6.31×10^1^ TCID_50_/mL)). Per decontamination method, nine PRCV-inoculated, decontaminated coupons (n=9) and three inoculated, decontaminated straps (n=3) were analysed in parallel to inoculated, untreated, positive control coupons (n=9) and straps (n=3). Mean log10 TCID_50_/mL and standard errors of the means are represented. P-values were computed by using a two-sided independent sample t-test, where ****P<0.0001, ***P<0.001, **P<0.01, *P<0.05, and ns is P≥0.05.

Following five cycles of SM UVGI, VHP exposure, and DH treatment, all MuNoV titres remained below the respective LOD of the assay, showing total loss of infectivity of over four orders of magnitude for UVGI and DH-treated coupons (4.65 and 4.29 log_10_ reduction, respectively), while titres of MuNoV recovered from VHP-treated coupons showed a loss of infectivity of almost four orders of magnitude (3.96 log_10_ reduction). Titres of MuNoV recovered from treated SM straps were reduced by 0.88, 2.39 (below the LOD), and 3.84 log_10_, respectively, post UVGI, VHP- and DH-treatment (Figure 9).

#### Multiple cycle decontamination of MuNoV-inoculated FFRs

Decontamination followed a similar pattern of MuNoV inactivation for FFR coupons decontaminated twice via DH, reducing viral titres by over three orders of magnitude (3.96 log_10_ reduction), and by over four orders of magnitude for VHP- and UVGI-treated FFR coupons (4.42, and 4.44 log_10_ reduction, respectively). UVGI-and DH-treatment of FFR straps reduced infectivity by 0.06 log_10_ (not significant), and 3.15 log_10_ (from 3.63 (±0.76) log_10_ TCID_50_/mL to below the LOD), respectively. Loss of infectivity could not be demonstrated subsequent to MuNoV elution from twice-VHP-treated FFR straps owing to poor virus recovery (Figure 10).

**Figure 10.**
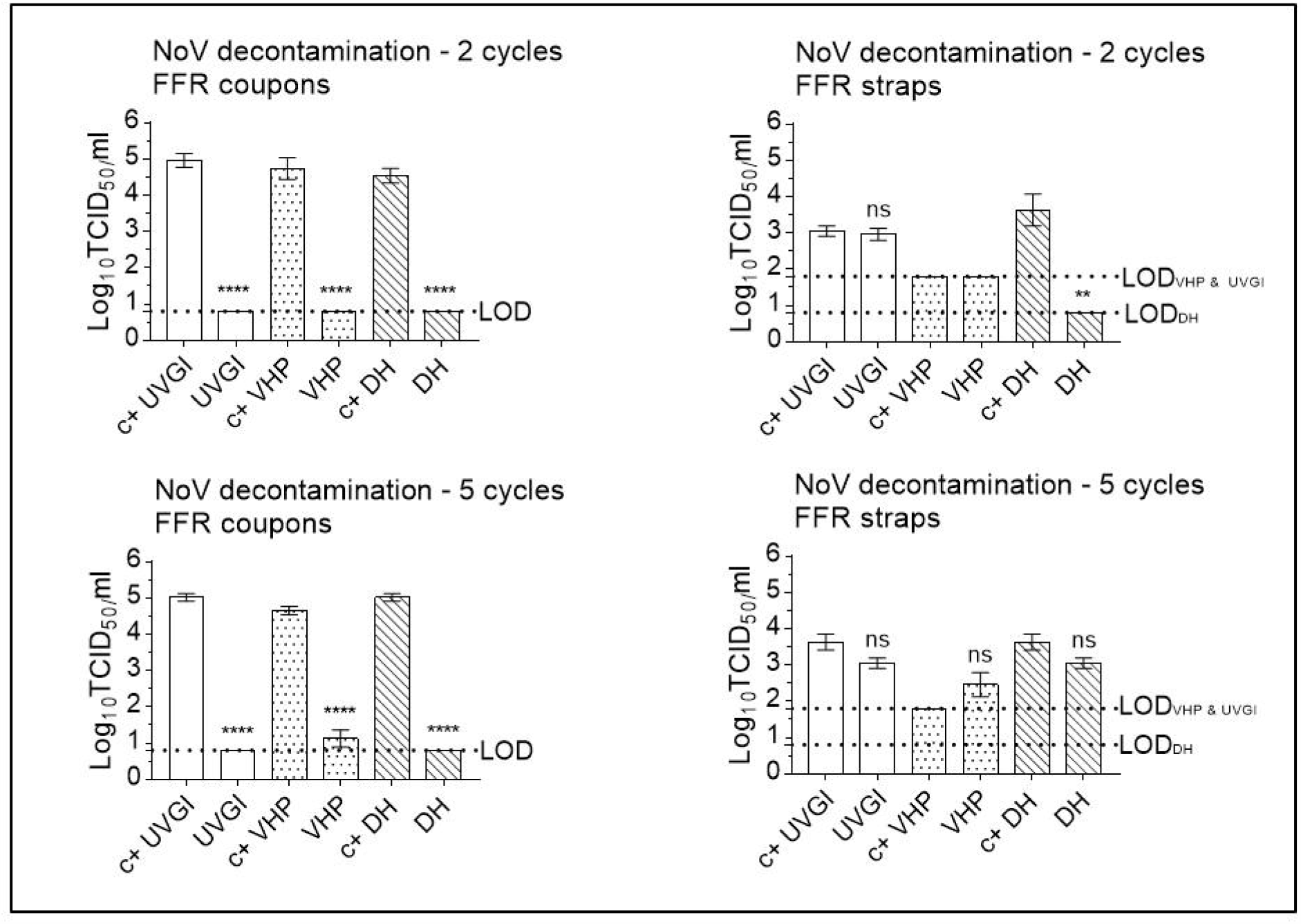
Murine norovirus (MuNoV) inactivation following multiple cycle filtering facepiece respirator (FFR) decontamination using dry heat (DH), vaporised hydrogen peroxide (VHP), and ultraviolet germicidal irradiation (UVGI). Titrations were performed after two or five (three in the case of DH) decontamination treatments on MuNoV-inoculated FFR coupons and straps. MuNoV infectivity was analysed in RAW264.7 cells. The cell culture limit of detection (LOD) was 0.80 log10 TCID_50_/mL (6.31×10^0^ TCID_50_/mL) for all analyses except those concerning VHP- and UVGI-treated FFR straps (1.80 log10 TCID_50_/mL (6.31×10^1^ TCID_50_/mL)). Per decontamination method, nine PRCV-inoculated, decontaminated coupons (n=9) and three inoculated, decontaminated straps (n=3) were analysed in parallel to inoculated, untreated, positive control coupons (n=9) and straps (n=3). Mean log10 TCID_50_/mL and standard errors of the means are represented. P-values were computed by using a two-sided independent sample t-test, where ****P<0.0001, ***P<0.001, **P<0.01, *P<0.05, and ns is P≥0.05.

Decontamination followed a similar pattern of MuNoV inactivation on FFR coupons after five iterations of UVGI, VHP, and DH treatments, reducing viral titres by over four orders of magnitude for UVGI- and DH-treated coupons (4.33 and 4.22 log_10_ reduction, respectively), and by less than three orders of magnitude for VHP-treated FFR coupons (2.84 log_10_ reduction). UVGI and DH-treatment of FFR straps reduced infectivity by less than one and over three orders of magnitude (0.65 (not significant) and 3.10 (not significant) log_10_ reduction, respectively); Loss of infectivity could not be demonstrated subsequent to MuNoV elution from VHP-treated FFR straps after five decontamination cycles owing to poor virus recovery (Figure 10).

## CONCLUSION

In conclusion, we showed that PRCV and MuNoV remain detectable on SMs and FFRs for up to five and seven days of passive decontamination at room temperature, necessitating either longer decontamination periods than currently recommended by the CDC or active decontamination techniques that can decontaminate PPE within a matter of hours. Three such active decontamination techniques were evaluated in this study with respect to their effect both on SM and FFR integrity and on the inactivation of the enveloped SARS-CoV-2 surrogate PRCV and non-enveloped human norovirus surrogate MuNoV. Single and multiple cycles of VHP-, UVGI-, and DH were shown to not adversely affect bacterial filtration efficiency of SMs. Single- and multiple UVGI did not adversely affect FFR filtration efficiency, while VHP and DH induced a slight decrease in FFR filtration efficiency after one or three decontamination cycles. Multiple cycles of VHP-, UVGI-, and DH decreased airflow resistance of SMs but did not adversely affect FFR breathability. All three active decontamination methods efficiently inactivated both viruses after five decontamination cycles, permitting demonstration of a loss of infectivity by more than three orders of magnitude. This multi-disciplinal, consolidated approach, wherein both SM and FFR integrity and the inactivation of a coronavirus and a hardier non-enveloped norovirus are investigated subsequent to multiple decontamination cycles thus provides important information on how often a given PPE item may be safely reused. The knowledge gained here will help close the existing gap between supply and demand and provide a multi-facetted measure of security to health-care personnel and the general public both during the Covid-19 pandemic and beyond, when established protocols for re-use of single-use only items may be upheld for environmental reasons.

## Data Availability

The authors confirm that the data supporting the findings of this study are available within the article [and/or] its supplementary materials.

## Non-standard abbreviations

FFR: filtering facepiece respirator
SM: surgical mask
SARS-CoV-2: severe acute respiratory syndrome coronavirus 2
PRCV: porcine respiratory coronavirus
MuNoV: murine norovirus
UVGI: ultraviolet germicidal irradiation
VHP: vaporised hydrogen peroxide
DH: dry heat
BFE: bacterial filtration efficiency

## ACKNOWLEDGEMENTS

The authors express their sincere gratitude to Amélie Matton and Frédéric de Meulemeester (AMB Ecosteryl, Mons, Belgium), Axel Kupisiewicz (LASEA, Sart-Tilman, Belgium), Pierre Leonard (Solwalfin, Belgium) for suggestions and technical and administrative support and thank Chantal Vanmaercke and Carine Boone for their excellent technical support.

## CONFLICTS OF INTEREST STATEMENT

The authors have no conflicts of interest to disclose.

## FUNDING SOURCE

This work was supported by a grant from the Walloon Region, Belgium (Project 2010053 -2020-“MASK -Decontamination and reuse of surgical masks and filtering facepiece respirators”) and the ULiège Fonds Spéciaux pour la Recherche 2020.

## SUPPLEMENTARY FIGURE CAPTIONS

**Supplementary Figure 1.**
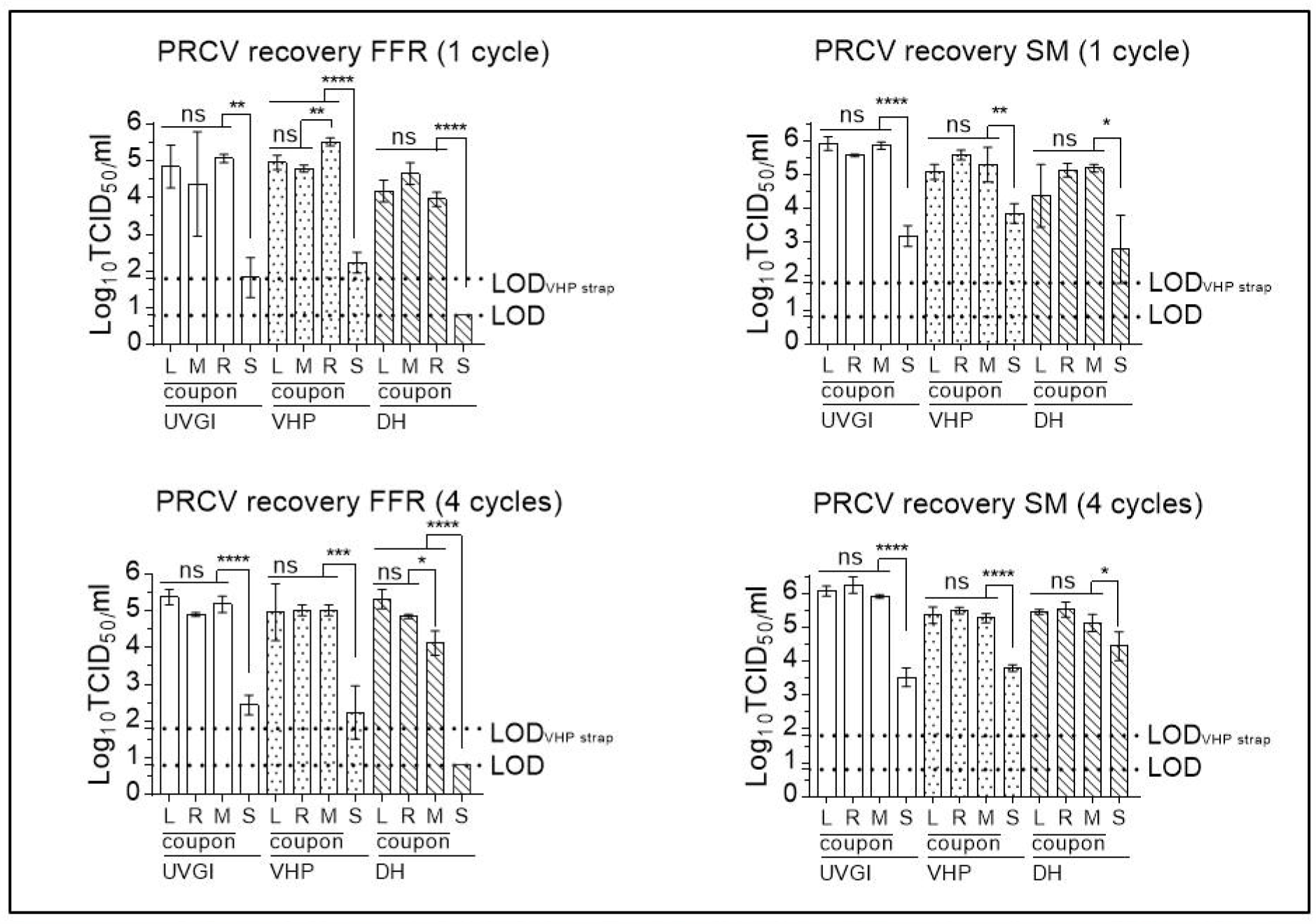
Recovery of porcine respiratory coronavirus (PRCV) after elution from filtering facepiece respirators (FFRs) and surgical masks (SMs) decontaminated either once or four times (twice in the case of DH assays) prior to virus inoculation. Infectious PRCV recovery was analysed in swine testicular cells. The cell culture limit of detection (LOD) was 0.80 log10 TCID_50_/mL (6.31×10^0^ TCID_50_/mL) for all analyses except those concerning VHP-treated SM or FFR straps (1.80 log10 TCID_50_/mL (6.31×10^1^ TCID_50_/mL)). Similar levels of virus recovery were detected for left, right and middle (L, R, M) (n=3) coupons of FFRs and SMs; recovery efficacy of infectious virus from straps (S) (n=3) deviated significantly in all analyses from the mean of all coupons and remained below the LOD for assays performed on DH-treated FFR straps. Mean log10 TCID_50_/mL and standard errors of the means are represented. P-values were computed by using a two-sided independent sample t-test to calculate differences between individual coupon values and differences between mean values of all coupons and straps, where ****P<0.0001, ***P<0.001, **P<0.01, *P<0.05, and ns.

**Supplementary Figure 2.**
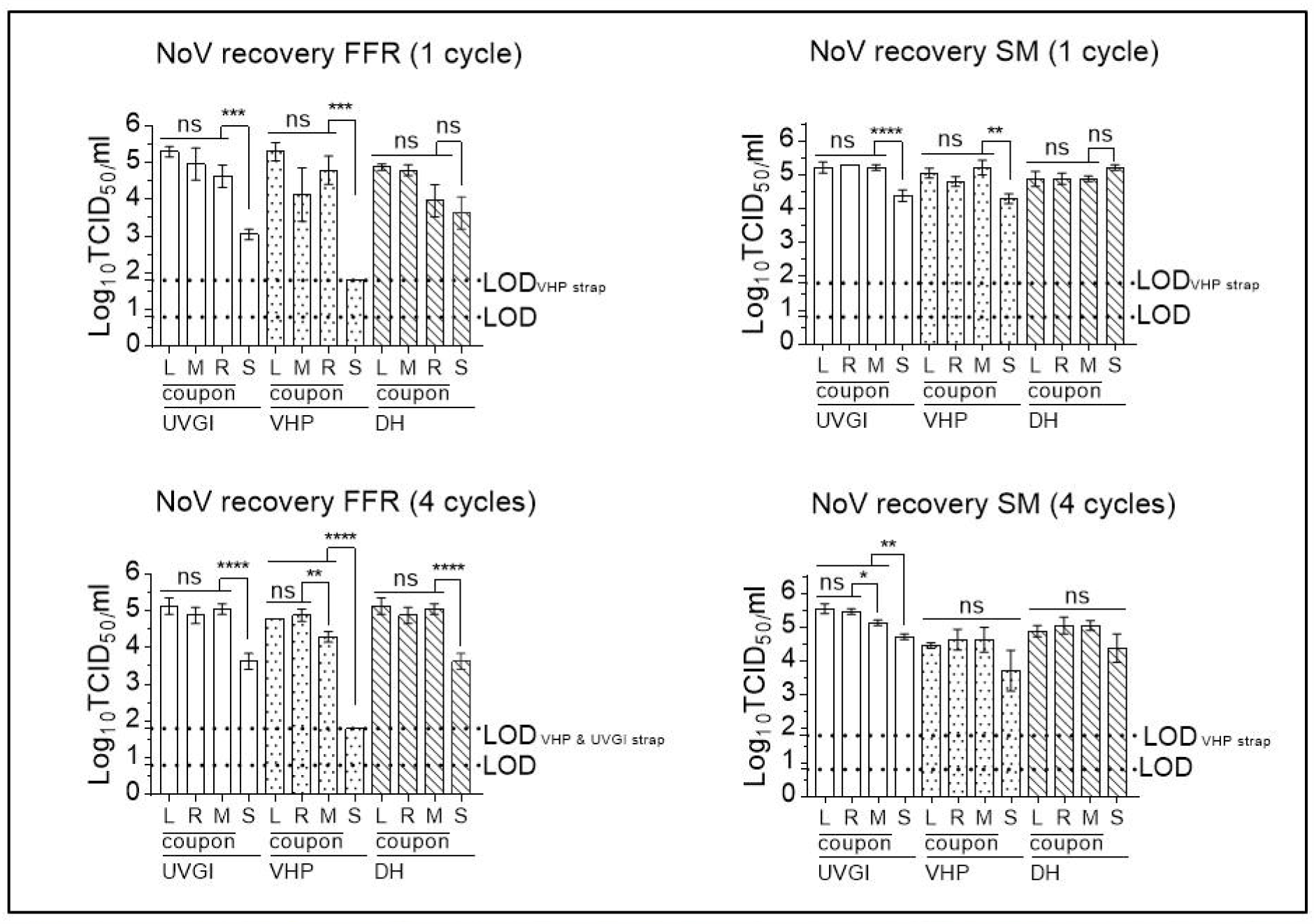
Recovery of murine norovirus (MuNoV) after elution from filtering facepiece respirators (FFRs) and surgical masks (SMs) decontaminated either once or four times (twice in the case of DH assays) prior to virus inoculation. Infectious MuNoV recovery was analysed in RAW264.7 cells. The cell culture limit of detection (LOD) was 0.80 log10 TCID_50_/mL (6.31×10^0^ TCID_50_/mL) for all analyses except those concerning VHP-treated SM- or FFR straps and UVGI- treated FFR straps (1.80 log10 TCID_50_/mL ((6.31×10^1^ TCID_50_/mL)). Similar levels of virus recovery were detected for left, right and middle (L, R, M) (n=3) coupons of FFRs and SMs; recovery efficacy of infectious virus from straps (S) (n=3) deviated significantly in all analyses from the mean of all coupons (except from DH-treated straps). Mean log10 TCID_50_/mL and standard errors of the means are represented. P-values were computed by using a two-sided independent sample t-test to calculate differences between individual coupon values and differences between mean values of all coupons and straps, where ****P<0.0001, ***P<0.001, **P<0.01, *P<0.05, and ns.

## Notes

### Competing Interest Statement

The authors have declared no competing interest.

### Author Declarations

No IRB and/or ethics committee approvals were neccessary in the context of this work.

